# Pre- and post-capillary exercise contributions to pulmonary hypertension in older adults

**DOI:** 10.1101/2025.01.21.25320932

**Authors:** Elizabeth Karvasarski, Joy Park, Simone Savaris, Anna Beale, Stephen P. Wright, Robert F. Bentley, John T. Granton, Susanna Mak

## Abstract

**Background:** Diagnosing pulmonary arterial hypertension (PAH) versus pulmonary hypertension associated with left hear disease (PH-LHD) can be challenging in patients with risk factors for both conditions. When resting pulmonary artery wedge pressure (PAWP) is proximate to a threshold of 15mmHg, exercise has been recommended to differentiate pre- versus post-capillary contributions to PH. To improve our understanding of this practice recommendation, we studied relationships between resting PAWP and the balance of pre- and post-capillary contributions to exercise PH.

**Methods:** Patients suspected of PAH (n=29, 72±2y, 52% F) with risk factors for LHD were prospectively recruited to undergo cycle ergometry at time of diagnostic right-heart catheterization. Hemodynamic data, including pressure-flow slopes and contributions of transpulmonary gradient (TPG) and PAWP to mPAP, were analyzed to evaluate pre- and post-capillary contributions, respectively, at rest and during exercise.

**Results:** PAWP ranged from 0 to 20 mmHg. Exercise pressure-flow slopes demonstrated 62% with post-capillary PH, and 31% with pre-capillary PH only. The relationship between resting PAWP and the pre- versus post-capillary contributions to exercise PH was not straightforward. Of patients with PAWP<12mmHg, 67% had post-capillary contributions to exercise PH. Conversely, 50% of patients with PAWP>15mmHg had pre-capillary contributions to exercise PH. Exercise-associated increases in pulmonary artery pressures were more strongly associated with pre-capillary contributions regardless of post-capillary contributions or the value of resting PAWP.

**Conclusion:** In this population, post-capillary contributions to exercise PH were commonly disclosed over a range of resting PAWP, including <12mmHg. The severity of exercise PH was determined by the pre-capillary contributions.

## INTRODUCTION

The diagnosis of pulmonary hypertension (PH) during right-heart catheterization (RHC) is based on a mean pulmonary artery pressure (mPAP) of >20mmHg; and the current approach to determining pre- versus post-capillary etiology is based on dichotomizing the supine resting pulmonary artery wedge pressure (PAWP) as ≤ or >15mmHg (Humbert et al., 2022). This approach may have limitations amongst PH patients with overlapping clinical risk factors for pre-capillary pulmonary arterial hypertension (PAH) and post-capillary PH associated with left-heart disease (PH-LHD). Exercise RHC has been recommended to improve phenotyping of PH for patients with left-heart risk factors (Humbert et al., 2022), particularly when the PAWP is “borderline”.

Definitions for a borderline or a “zone of uncertainty” (Rayner et al., 2024) have been proposed as between 13 to 15mmHg (Humbert et al., 2022) or between 12 to 18mHg (Rayner et al., 2024), in part based on the demonstration that the upper limit of normal for PAWP is approximately 12mmHg (Zeder et al., 2024). Studies conducted in healthy subjects (Esfandiari et al., 2019; Zeder et al., 2022) have also clarified that the upper limit of normal for the increases in mPAP responses relative to increases in cardiac output (CO) during exercise (ΔmPAP/ΔCO) is 3WU, which now defines Exercise PH (Humbert et al., 2022).

However, mPAP reflects the summation of both pre- and post-capillary responses and the hemodynamic criteria that distinguish these contributions to Exercise PH are not fully clear. We have shown that in healthy adults, increases in mPAP during exercise are closely related to changes in PAWP, the post-capillary contribution (Esfandiari et al., 2017) and there is some consensus that the upper limit of normal for exercise ΔPAWP/ΔCO is 2WU (Eisman et al., 2018). There is less consensus regarding measurements that define the range of normal and abnormal pre-capillary responses to exercise (Zeder et al., 2022). Changes in the transpulmonary gradient (TPG: mPAP – PAWP) relative to CO may reflect the pre-capillary component. We have further shown that in health, exercise-associated pre-capillary responses include declines in calculate pulmonary vascular resistance (PVR), but also declines in PA compliance (PAC) that appear related to changes in PAWP (Bentley et al., 2020; Wright, Granton, et al., 2016).

In this study, we tested the assumption that resting supine PAWP would be related to the balance of pre- and post-capillary contributions to exercise mPAP. We examined pressure-flow relationships and the interaction of PVR, PAC, and PAWP during exercise, as well as the value of resting PAWP to predict the pre- versus post-capillary contributions to exercise mPAP. Study patients were prospectively recruited based on a high suspicion of PAH with an intermediate to high clinical likelihood of PH-LHD, the target population with indications for exercise RHC. Patterns of hemodynamic exercise responses were contrasted to historical healthy controls.

## METHODS

### Older Patients with Suspected PH

Our PH center conducted a prospective observational cohort study of 43 patients suspected of PAH but with ≥1 cardiovascular risk factor referred from the Pulmonary Hypertension Program, University of Toronto. Patients were recruited to undergo exercise at the time of their first diagnostic RHC performed at the Cardiac Catheterization Clinical Research Laboratory at Sinai Health.

The inclusion criteria were: 1) >45 years; 2) ≥1 of BMI >30kg/m^2^, hypertension treated with >1 medication, diabetes mellitus treated with oral hypoglycemic agents or insulin, documented coronary artery disease, or atrial fibrillation; 3) 2-dimensional echocardiogram documenting both left ventricular ejection fraction >55%; and peak velocity of the tricuspid regurgitation jet >2.8m/s consistent with at least moderate probability of PH (Humbert et al., 2022). Major exclusions were: 1) previous diagnosis of PAH confirmed by RHC; 2) previously received PAH therapy; 3) documented chronic obstructive pulmonary disease, hypoxic lung disease, or known thromboembolic pulmonary disease (overtly suspected Group 3/4 PH); 4) admission for congestive heart failure within 6 months; 5) structural cardiac valvular disease or intracardiac shunting; 6) infiltrative or hypertrophic cardiomyopathy, or pericardial disease; 7) recent (<6months) acute coronary syndrome, myocardial infarction, or unstable ventricular arrhythmia. Institutional research ethics boards at Mount Sinai Hospital (#18-0257-A) and the University Health Network (19-5069.0) approved this study.

For the current analysis, we excluded patients that did not complete exercise, patients with marked elevation of resting PAWP (>22mmHg), and those with increases in CO<1L/min with exercise as calculations of pressure-flow slopes became confounded.

### Healthy Subjects

We previously conducted exercise hemodynamic investigations to compare responses between men and women (n=36, 50% female) (Bentley et al., 2020; Esfandiari et al., 2017; Wright, Esfandiari, et al., 2016; Wright, Granton, et al., 2016). These subjects were recruited from the community by media advertising and had no history of cardiac or systemic disease.

For healthy subjects, hemodynamics and demographic characteristics have been reported; however, a detailed analysis of pre- and post-capillary hemodynamic responses were not.

### Catheterization Procedures

RHC was performed and hemodynamics were recorded at rest and during exercise as previously published by our laboratory (Karvasarski et al., 2023). Supine hemodynamics were recorded first and pressures sampled from the right atrium (RA), right ventricle (RV), and PA. PAWP was measured by intermittent inflation of the balloon-tip. Thermodilution CO was measured in triplicate (≤10% variation). Patients were transferred to cycle ergometer and tilted into a semi-upright position. Transducers were re-zeroed at the level of the midaxillary line. Hemodynamics were resampled in the semi-upright position (control) prior to initiating a 1-minute warm-up with unloaded pedaling. The workload was determined based on the Medical Research Council (MRC) dyspnea scale determined prior to the study and one work-rate was selected based on the MRC score: MRC >3: 15W, MRC <3: 25W for women or 40W for men.

### Hemodynamic Analysis

Hemodynamics reported included heart rate (HR), systemic blood pressure, mean RA pressure (mRAP), systolic/diastolic/mean PA pressure (PASP/PADP/mPAP), CO, and mean PAWP. Additional calculated variables included the transpulmonary gradient (TPG)=mPAP– PAWP, diastolic pressure gradient (DPG)=PADP–PAWP, PVR=TPG/CO, PA pulse pressure (PP)=PASP–PADP, PAC=stroke volume (SV)/PP, and resistance-compliance (RC) time (s)=[TPG/(HRxPP)]x60.

### Resting Classifications and PAWP Groupings

Resting hemodynamic classifications for PH, PAH and PH-LHD were made as per guideline recommendations (Humbert et al., 2022). To evaluate the relationship between resting PAWP and subsequent exercise hemodynamics, we considered PAWP Normal if <12mmHg, Borderline if 12-15mmHg, and High if >15mmHg (Humbert et al., 2022; Rayner et al., 2024; Zeder et al., 2022). Recognizing these groupings are arbitrary, we also evaluated PAWP as a continuous variable.

### Definitions of Pre- and Post-Capillary Hemodynamics: Pressure-Flow Relationships with Exercise

Exercise PH was defined as per the 2022 ESC/ERS guidelines: change in mPAP adjusted for the change in CO (ΔmPAP/ΔCO) >3WU (Humbert et al., 2022).

Pre- and post-capillary contributions to mPAP at rest and with exercise were considered according to the following equations:

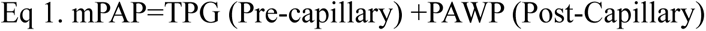

The change in mPAP with exercise can be represented as

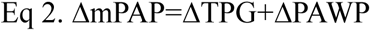

The absolute value of the mPAP with exercise can be represented as

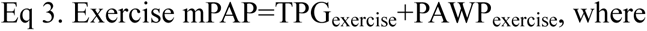

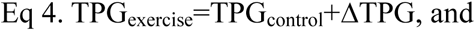

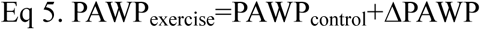

The ΔmPAP/ΔCO slope can also be written as

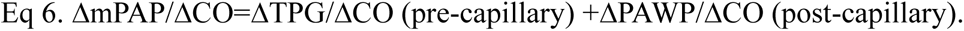

The variables mPAP and PAWP were measured via RHC, TPG was calculated based on the equation above, and CO was measured via thermodilution.

### Exercise Hemodynamic Groupings

Exercise PH classifications were based on pressure flow relationships:

No Exercise PH: ΔmPAP/ΔCO ≤3WU
Exercise PH: ΔmPAP/ΔCO >3WU
Exercise PAH: ΔmPAP/ΔCO >3WU AND ΔPAWP/ΔCO ≤2WU
Exercise PH-LHD: ΔmPAP/ΔCO >3WU and ΔPAWP/ΔCO >2WU

Exercise PH-LHD_isolated_: ΔTPG/ΔCO ≤2WU
Exercise PH-LHD_combined_: ΔTPG/ΔCO >2WU

ΔPAWP/ΔCO represents the post-capillary contributor versus ΔTPG/ΔCO, the pre-capillary contributor to PH as above (Ho et al., 2020; Zeder et al., 2022).

We considered the upper limit of normal for ΔPAWP/ΔCO is 2WU and for ΔTPG/ΔCO is 2WU (Eisman et al., 2018; Esfandiari et al., 2017; Zeder et al., 2022).

### Statistical Analysis

Statistics were completed using SPSS 29.0 (IBM Corp, Armonk, NY), R 4.3.3, and Prism 10 (GraphPad Software, San Diego, CA). Normality was assessed quantitatively with a Shapiro– Wilk test. Continuous data was presented as mean ± standard deviation (SD) if normally distributed and as median [quartile 1 (Q1) – quartile 3 (Q3)] if not normally distributed. Between group comparisons were conducted with a t-test or analysis of variance (ANOVA) with Bonferroni correction in the case of normally distributed data and a Mann-Whitney U or Kruskal-Wallis test if data were not normally distributed. Chi-squared analyses were performed for categorical variables. Hemodynamic parameters measured at control and exercise were analyzed using a two-way repeated measures ANOVA, with the stage of the procedure as one factor (including control and exercise) and group as the second factor. Associations between continuous variables were explored using multiple linear regression. Receiver operator curves were generated to determine the ability of resting hemodynamics to predict ΔPAWP/ΔCO >2WU, and ΔTPG/ΔCO >2WU. An area under the curve was calculated for each, and if significant, the Youden index was used to determine the value that achieved maximal sensitivity and specificity. An α level of 0.05 was considered statistically significant.

## RESULTS

### Patient Characteristics and Baseline Hemodynamics

This analysis comprised 29 patients after exclusions. Although recruited on the basis of a high likelihood of PH, 22 had incident PH and 7 did not have PH at rest (No PH_rest_). Table 1 demonstrates the patient characteristics and baseline hemodynamics. The population had a mean age of 72±9 years, 47% were women, and the median H2FPEF score was 6.

**Table 1.** Older PH patient characteristics for the PAWP groupings.

|  | **All Older PH Patients**<br>**n=29** | **“Normal” Zone**<br>**PAWP**<br>**<12mmHg**<br>**n=18** | **“Borderline” Zone**<br>**PAWP**<br>**12-15mmHg**<br>**n=5** | **“High” Zone**<br>**PAWP**<br>**>15mmHg**<br>**n=6** |
| --- | --- | --- | --- | --- |
| *Characteristics* |  |  |  |  |
| Age, (y) | 72 ± 10 | 69 ± 11 | 78 ± 4 | 77 ± 7 |
| Women, n (%) | 15 (52) | 12 (67) | 2 (40) | 1 (17) |
| Weight (kg) | 78.4 ± 15.2 | 79.6 ± 15.5 | 67.3 ± 14.1 | 84.3 ± 12.0 |
| Height (m) | 1.7 ± 0.1 | 1.7 ± 0.1 | 1.7 ± 0.1 | 1.7 ± 0.1 |
| BSA (m<sup>2</sup>) | 1.9 ± 0.2 | 1.9 ± 0.2 | 1.8 ± 0.2 | 2.0 ± 0.1 |
| BMI (kg/m<sup>2</sup>) | 28.0 ± 5.2 | 29.2 ± 5.2 | 23.2 ± 3.2 | 28.4 ± 4.4 |
| BMI >30kg/m<sup>2</sup>, n (%) | 10 (34) | 8 (44) | 0 | 2 (33) |
| Hypertension | 24 (83) | 13 (72) | 5 (100) | 6 (100) |
| Diabetes | 10 (34) | 6 (33) | 2 (40) | 2 (33) |
| Previous CAD | 4 (14) | 1 (6) | 1 (20) | 2 (33) |
| Atrial fibrillation | 15 (52) | 5 (28) | 4 (80)* | 6 (100)* |
| H2FPEF Score | 6 (3-7) | 4 (3-6) | 7 (5-7) | 8 (7-8) |
| Group 1 PH associations, n (%) | 9 (31) | 8 (44) | 0 | 1 (17) |
| No PH<sub>rest</sub> | 7 (24) | 6 (33) | 1 (20) | 0 |
| PAH<sub>rest</sub> | 16 (55) | 12 (67) | 4 (80) | 0 |
| PH-LHD<sub>rest</sub> | 6 (21) | 0 | 0 | 6 (100) |
| *Baseline Hemodynamics* |  |  |  |  |
| HR (bpm) | 71 ± 14 | 76 ± 12 | 63 ± 18 | 61 ± 12 |
| MAP (mmHg) | 103 ± 16 | 101 ± 15 | 99 ± 8 | 113 ± 18 |
| SV (mL) | 74 ± 22 | 71 ± 19 | 71 ± 16 | 85 ± 32 |
| CO (L/min) | 5.0 ± 1.2 | 5.3 ± 1.3 | 4.3 ± 0.9 | 4.9 ± 1.2 |
| RAP (mmHg) | 3 ± 3 | 2 ± 2 | 5 ± 3 | 8 ± 3 |
| mPAP | 28 ± 11 | 25 ± 11 | 27 ± 7 | 39 ± 9 |
| PAWP | 9 ± 6 | 6 ± 4 | 13 ± 1 | 17 ± 2 |
| PVR (WU) | 3.7 (2.1-5.8) | 3.5 (2.0-5.9) | 2.4 (2.1-5.2) | 4.8 (3.5-6.6) |
| PAC (mL/mmHg) | 2.0 (1.5-3.1) | 2.3 (1.9-3.3) | 2.3 (1.6-2.8) | 1.5 (1.2-2.5) |
| RC time (s) | 0.5 ± 0.1 | 0.5 ± 0.1 | 0.4 ± 0.1 | 0.4 ± 0.1 |

The range of resting PAWP among the study population was 0 to 20mmHg. At supine rest, 62% demonstrated PAWP in the Normal Zone (PAWP <12mmHg), 17% in the Borderline Zone (12-15mmHg), and 21% in the High Zone (PAWP >15mmHg). Table 1 shows clinical characteristics based on PAWP groupings and tertiles. Patients in the Borderline and High PAWP Zone groups were more likely to have systemic hypertension and atrial fibrillation.

### Patient Classifications based on Resting PAWP and ΔPAWP/ΔCO

Hemodynamic data relevant to pressure-flow slopes and exercise classifications for PAWP groupings are included in Table 2. Overall, Exercise PH was observed in 27of 29 older patients with suspected PH; a ΔmPAP/ΔCO slope <3WU was observed in only 2 of the No PH_rest_ patients.

**Table 2.** Exercise and delta hemodynamics in PAWP groupings in the older PH patients.

|  | Normal Zone<br>PAWP <12mmHg<br>n=18 |  | Borderline Zone<br>PAWP 12-15mmHg<br>n=5 |  | High Zone<br>PAWP >15mmHg<br>n=6 |  |
| --- | --- | --- | --- | --- | --- | --- |
|  | Ex | Δ | Ex | Δ | Ex | Δ |
| **CO (L/min)** | 8.1 ± 2.2 | 2.9 ± 1.1 | 6.5 ± 0.6 | 2.6 ± 0.6 | 6.9 ± 2.6 | 2.3 ± 1.8 |
| **SV (mL)** | 81 ± 21 | 11 ± 8 | 76 ± 44 | 18 ± 30 | 81 ± 3 | 0 ± 22 |
| **mPAP (mmHg)** | 41 ± 18 | 18 ± 8 | 46 ± 15 | 20 ± 9 | 54 ± 11 | 18 ± 6 |
| **PAWP** | 13 ± 7 | 8 ± 6 | 19 ± 6 | 7 ± 9 | 21 ± 6* | 7 ± 8 |
| **TPG** | 28 ± 16 | 10 ± 7 | 26 ± 15 | 13 ± 6 | 32 ± 14 | 11 ± 8 |
| **ΔmPAP/ΔCO (WU)** | 6.25 (3.71-12.11) |  | 6.92 (4.49-10.29) |  | 7.41 (5.69-19.56) |  |
| **ΔPAWP/ΔCO** | 2.42 (0.86-5.08) |  | 3.06 (-1.12-4.73) |  | 2.80 (-0.10-7.99) |  |
| **ΔTPG/ΔCO** | 3.19 (1.50-5.38) |  | 3.47 (2.95-8.41) |  | 7.11 (0.25-13.24) |  |
| **Exercise PH, n (% of Zone)** | 16 (89) |  | 5 (100) |  | 6 (100) |  |
| **Exercise PAH/PH-LHD** | 4/12 (22/67) |  | 2/3 (40/60) |  | 3/3 (50/50) |  |
| **Exercise PH-LHD<sub>isolated/combined</sub>** | 4/8 (22/44) |  | 0/3 (0/60) |  | 2/1 (33/17) |  |

Data represented as mean ± SD if normally distributed or median (quartile 1 – quartile 3) if not normally distributed. \* P<0.05 vs Normal Zone. Exercise PH: ΔmPAP/ΔCO >3WU, Exercise PAH: ΔmPAP/ΔCO >3WU and ΔPAWP/ΔCO ≤2WU, Exercise PH-LHD: ΔmPAP/ΔCO >3WU, ΔPAWP/ΔCO >2WU, Exercise PH-LHD<sub>isolated</sub>: ΔTPG/ΔCO ≤2WU, Exercise PH-LHD<sub>combined</sub>: ΔTPG/ΔCO >2WU.

Exercise PH-LHD (ΔPAWP/ΔCO >2WU) was observed in 62% of our recruited patients. Values for the ΔPAWP/ΔCO pressure-flow slopes based on resting PAWP groupings are illustrated in Figure 1A. We did not observe a correlation between resting PAWP and ΔPAWP/ΔCO slope. The distributions of exercise classifications based on PAWP groupings is illustrated in Figure 1B. Exercise PH-LHD was still the predominant classification (67%) amongst the 18 patients in the PAWP ‘Normal Zone’. Overall, Exercise PAH was observed less commonly in 31%. Patients with exercise PAH were detected across all PAWP groupings, even amongst patients in the PAWP ‘High Zone’.

**Figure 1.**
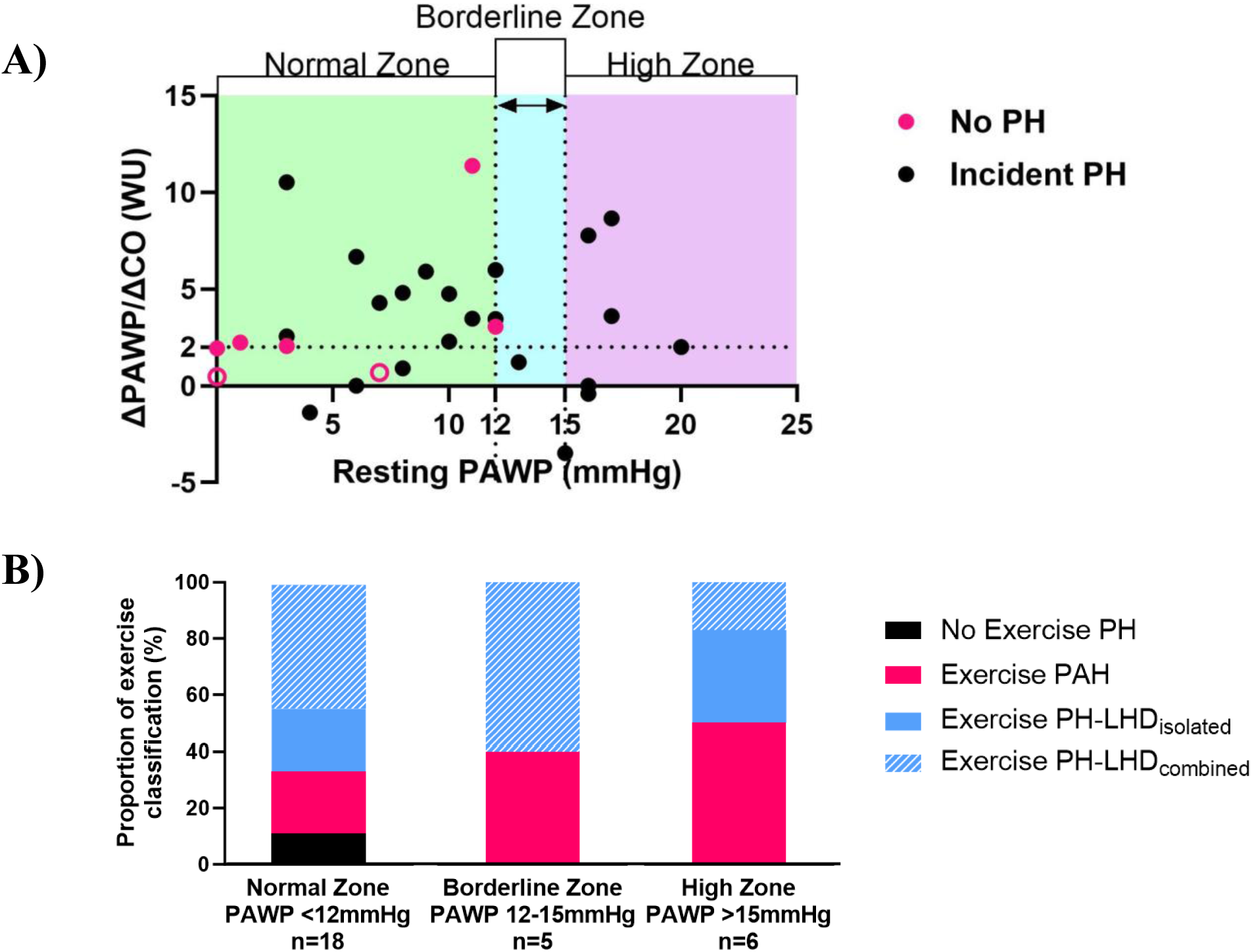
Resting PAWP against the ΔPAWP/ΔCO slope in the No PH_rest_, and Incident PH patients and Proportion of patients in PH groupings. A: Black circles represent incident PH, pink circles represent older PH patients with No PH_rest_, and open pink circles demonstrate patients with No PH at rest and exercise. The ‘Borderline’ PAWP zone are also illustrated. B: Black bars represent patients with No Exercise PH (ΔmPAP/ΔCO ≤3WU); Pink bars represent Exercise PAH (ΔmPAP/ΔCO >3WU and ΔPAWP/ΔCO ≤2WU); Blue bars represent Exercise PH-LHD (ΔmPAP/ΔCO >3WU, ΔPAWP/ΔCO >2WU); Blue solid bars represent Exercise PH-LHD_isolated_ (ΔTPG/ΔCO ≤2WU); Blue hatched bars represent Exercise PH-LHD_combined_ (ΔTPG/ΔCO >2WU).

### Patterns of Pre- and Post-capillary Contributions to Rest and Exercise PA Pressure: Contrasts to Normal Physiology

The summed contributions of PAWP (post-capillary) and TPG (pre-capillary) to resultant rest and exercise mPAP across healthy subjects and older PH patients are illustrated in Figure 2, and the patterns of association between PAWP and TPG with resultant mPAP are illustrated in Figure 3.

**Figure 2.**
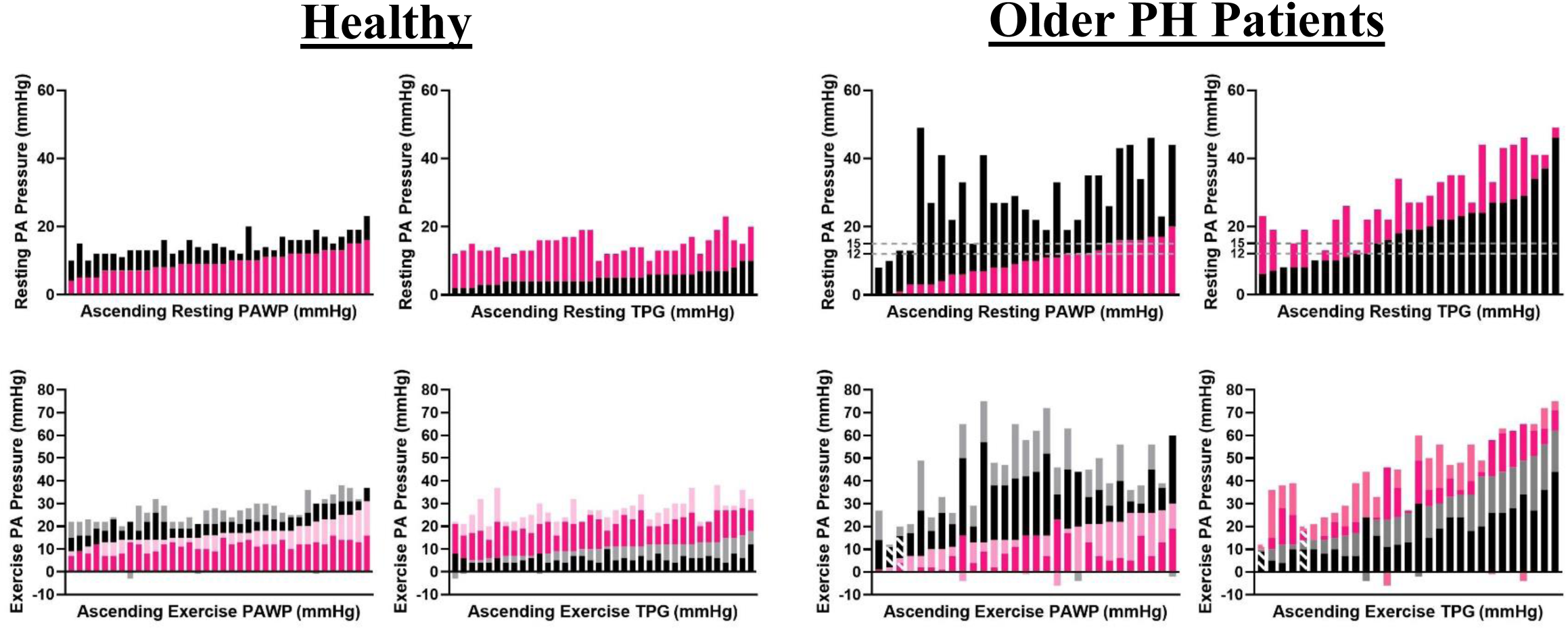
mPAP = TPG + PAWP for Healthy and Older PH patients at rest and exercise. Top Row illustrates supine rest condition: Resting mPAP = resting TPG (black bars) + resting PAWP (pink bars). Graphs are arranged by ascending resting PAWP or TPG as indicated on the x-axis. Bottom row illustrates exercise condition: Exercise mPAP = exercise TPG + exercise PAWP, where exercise TPG = control TPG (black bars) + ΔTPG (grey bars) and exercise PAWP = control PAWP (pink bars) + ΔPAWP (light pink bars). ΔTPG exercise TPG – control TPG, and ΔPAWP = exercise PAWP – control PAWP. Hatched bars indicate patients with No PH at rest and No exercise PH.

**Figure 3.**
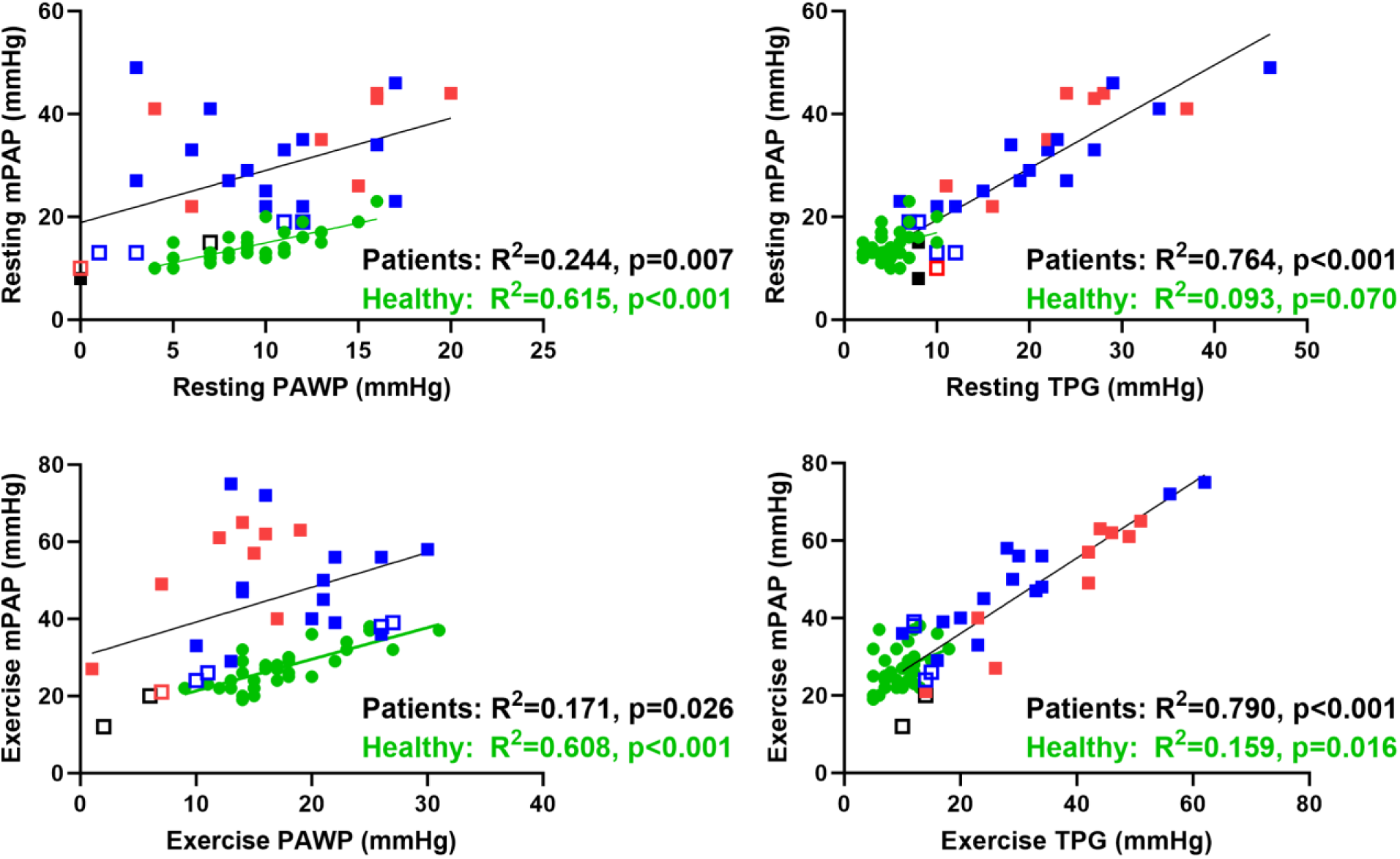
Linear relationship between PAWP, TPG, and mPAP in Healthy subjects and Older PH patients at rest and exercise. Healthy subjects indicated by green circle and green linear regression line. Patients indicated by black linear regression line, red squares indicate Exercise PAH, blue squares indicate Exercise PH-LHD, open symbols indicate No PH_rest_, and black open squares indicate No PH at rest and exercise.

In healthy subjects, mPAP is more strongly associated with post-capillary contributions both at rest and exercise (Table 3). At rest, mPAP was significantly related to PAWP (R^2^=0.615, p<0.001), but not TPG (R^2^=0.093, p=0.070). Exercise mPAP was also more strongly related to exercise PAWP (R^2^=0.608, p<0.001), and more modestly to TPG (R^2^=0.159, p=0.016). Amongst the older PH patient population, we observed a distinctly contrasting pattern such that mPAP was more strongly associated with pre-capillary contributions both at rest and exercise. At rest mPAP was modestly related to PAWP (R^2^=0.244, p=0.007); and more strongly to TPG (R^2^=0.764, p<0.001). Similarly exercise mPAP was strongly related to exercise TPG (R^2^=0.790, p<0.001), and less so to exercise PAWP (R^2^=0.171, p=0.026). Stronger relationships between mPAP and pre-capillary contributions, both at rest and exercise, were observed even when separately considering patients classified as Exercise PAH or PH-LHD.

**Table 3.** Linear regression analysis for Healthy, older PH patients, and the exercise PH subgroups between PAWP, TPG, and mPAP.

|  |  | **Healthy**<br>**n=36** | **Older PH**<br>**patients**<br>**n=29** | **Ex PAH**<br>**n=9** | **Ex PH-LHD**<br>**n=18** |
| --- | --- | --- | --- | --- | --- |
| **Resting mPAP** |  |  |  |  |  |
| **Resting PAWP (mmHg)** | **R<sup>2</sup>** | **0.615** | **0.244** | **0.498** | 0.033 |
|  | **P** | **<0.001** | **0.007** | **0.034** | 0.471 |
| **Resting TPG** | **R<sup>2</sup>** | 0.093 | **0.764** | **0.702** | **0.795** |
|  | **P** | 0.070 | **<0.001** | **0.005** | **<0.001** |
| **Exercise mPAP** |  |  |  |  |  |
| **Exercise PAWP (mmHg)** | **R<sup>2</sup>** | **0.608** | **0.171** | **0.495** | 0.046 |
|  | **P** | **<0.001** | **0.026** | **0.035** | 0.394 |
| **Exercise TPG** | **R<sup>2</sup>** | **0.159** | **0.790** | **0.899** | **0.807** |
|  | **P** | **0.016** | **<0.001** | **<0.001** | **<0.001** |

P: p-value.

### Hemodynamic Responses of Patients Classified as Exercise PAH or Exercise PH-LHD

Hemodynamic responses observed in patients classified as Exercise PAH or Exercise PH-LHD are shown in Table 4. At control, prior to exercise, there were no statistically significant differences. With exercise, CO and SV were also not different between Exercise PAH and Exercise PH-LHD. As such, the pressure-flow slope classifications were then derived from larger PAWP and ΔPAWP responses in the Exercise PH-LHD group, and larger TPG and DPG responses in the Exercise PAH group. When comparing subgroups of Exercise PH-LHD_isolated_ and PH-LHD_combined_ (also shown in Table 4), there were notably no differences in control PAWP, exercise PAWP or ΔPAWP values. The main differences were exercise changes in PA PP, TPG, PASP, and PVR, which were all significantly greater in the Exercise PH-LHD_combined_ compared to Exercise PH-LHD_isolated_ groups.

**Table 4.** Control, exercise, and delta hemodynamics in the patient exercise groupings.

|  | Exercise PAH n=9 |  |  | Exercise PH-LHD n=18 |  |  |
| --- | --- | --- | --- | --- | --- | --- |
|  | Control | Exercise | Δ | Control | Exercise | Δ |
| HR (bpm) | 71 ± 14 | 105 ± 24 | 34 ± 18 | 70 ± 15 | 97 ± 27 | 27 ± 20 |
| SV (mL) | 71 ± 21 | 78 ± 28 | 7 ± 14 | 67 ± 19 | 78 ± 27 | 11 ± 19 |
| CO (L/min) | 4.84 ± 1.14 | 7.69 ± 2.28 | 2.84 ± 1.29 | 4.52 ± 0.83 | 7.07 ± 1.65 | 2.55 ± 1.19 |
| RAP (mmHg) | 3 ± 4 | 12 ± 8 | 8 ± 4 | 0 ± 3 | 8 ± 4 | 7 ± 4 |
| PASP | 58 ± 26 | 79 ± 25 | 22 ± 13 | 42 ± 21 | 71 ± 25 | 31 ± 13 |
| PADP | 18 ± 10 | 28 ± 11 | 10 ± 6 | 13 ± 8 | 26 ± 9 | 14 ± 6 |
| mPAP | 33 ± 14 | 49 ± 16 | 16 ± 7 | 25 ± 13 | 45 ± 14 | 22 ± 7 |
| PP | 40 ± 18 | 51 ± 16 | 12 ± 10 | 29 ± 15 | 46 ± 19 | 17 ± 15 |
| PAWP | 11 ± 8 | 12 ± 6 | 1 ± 4 | 7 ± 5 | 19 ± 6* | 13 ± 5* |
| TPG | 22 ± 9 | 37 ± 13 | 16 ± 6 | 17 ± 11 | 26 ± 14 | 10 ± 7* |
| DPG | 6 ± 5 | 16 ± 8 | 10 ± 5 | 6 ± 7 | 7 ± 8* | 1 ± 5* |
| PVR (WU) | 5.13 (2.41-6.43) | 5.58 (3.65-7.00) | 0.48 (0.31-1.24) | 3.28 (1.74-5.47) | 3.52 (1.98-4.79) | 0.09 (-0.75-0.65) |
| PAC (mL/mmHg) | 1.69 (1.16-3.46) | 1.33 (1.07-2.42) | -0.41 (-1.02- -0.15) | 2.44 (1.70-3.35) | 1.73 (1.21-2.69) | -0.49 (-1.45- -0.20) |
| RC time (s) | 0.49 ± 0.11 | 0.43 ± 0.10 | -0.06 ± 0.07 | 0.50 ± 0.15 | 0.36 ± 0.11 | -0.17 ± 0.13 |
|  | Exercise PH-LHD<sub>isolated</sub> n=6 |  |  | Exercise PH-LHD<sub>combined</sub> n=12 |  |  |
|  | Control | Exercise | Δ | Control | Exercise | Δ |
| HR (bpm) | 67 ± 11 | 102 ± 20 | 35 ± 23 | 72 ± 17 | 95 ± 30 | 23 ± 19 |
| SV (mL) | 73 ± 22 | 76 ± 20 | 3 ± 13 | 64 ± 17 | 80 ± 31 | 15 ± 21 |
| CO (L/min) | 4.70 ± 0.71 | 7.66 ± 2.36 | 2.96 ± 1.82 | 4.43 ± 0.90 | 6.78 ± 1.17 | 2.35 ± 0.74 |
| RAP (mmHg) | 1 ± 5 | 6 ± 6 | 5 ± 4 | 0 ± 2 | 8 ± 4 | 8 ± 3 |
| PASP | 39 ± 24 | 59 ± 15 | 20 ± 11 | 44 ± 21 | 77 ± 27 | 34 ± 11‡ |
| PADP | 11 ± 8 | 24 ± 9 | 13 ± 4 | 15 ± 8 | 27 ± 9 | 12 ± 7 |
| mPAP | 22 ± 4 | 38 ± 11 | 16 ± 6 | 26 ± 12 | 48 ± 15 | 23 ± 7 |
| PP | 29 ± 16 | 36 ± 9 | 7 ± 11 | 29 ± 15 | 51 ± 12 | 22 ± 11‡ |
| PAWP | 7 ± 8 | 21 ± 9 | 14 ± 6 | 8 ± 4 | 18 ± 5 | 11 ± 5 |
| TPG | 16 ± 10 | 18 ± 7 | 2 ± 5 | 18 ± 12 | 30 ± 16 | 12 ± 4‡ |
| DPG | 4 ± 6 | 3 ± 7 | -1 ± 5 | 7 ± 8 | 8 ± 9 | 1 ± 4 |
| PVR (WU) | 3.19 (1.79-4.89) | 2.62 (1.78-3.03) | -0.57 (-1.47- -0.10) | 3.65 (2.12-4.94) | 4.30 (2.59-5.23) | 0.53 (0.12-0.66)‡ |
| PAC (mL/mmHg) | 2.47 (1.90-4.58) | 2.04 (1.52-3.29) | -0.57 (-1.29- -0.17) | 2.40 (1.63-3.14) | 1.68 (1.21-2.44) | -0.49 (-1.45- -0.17) |
| RC time (s) | 0.47 ± 0.15 | 0.30 ± 0.07 | -0.17 ± 0.13 | 0.51 ± 0.15 | 0.39 ± 0.11 | -0.12 ± 0.14 |

Data represented as mean ± SD if normally distributed or median (quartile 1 – quartile 3) if not normally distributed. \*P<0.05 vs Exercise PAH, ‡P<0.05 vs Exercise PH-LHD<sub>isolated</sub>.

Among the patient population, we further examined the behavior of the pre-capillary circulation during exercise by evaluating PVR, PAC, and RC time. Overall, PAC declined significantly, and RC time shortened, with no change in PVR, but there were contrasts observed between exercise classifications (Figure 4). Among the Exercise PAH subgroup, PAC, PVR and RC time did not change significantly; in contrast among the Exercise PH-LHD subgroup, PAC declined, and RC time shortened significantly with exercise. This behavior was observed in both Exercise PH-LHD_combined_ and Exercise PH-LHD_isolated_.

**Figure 4.**
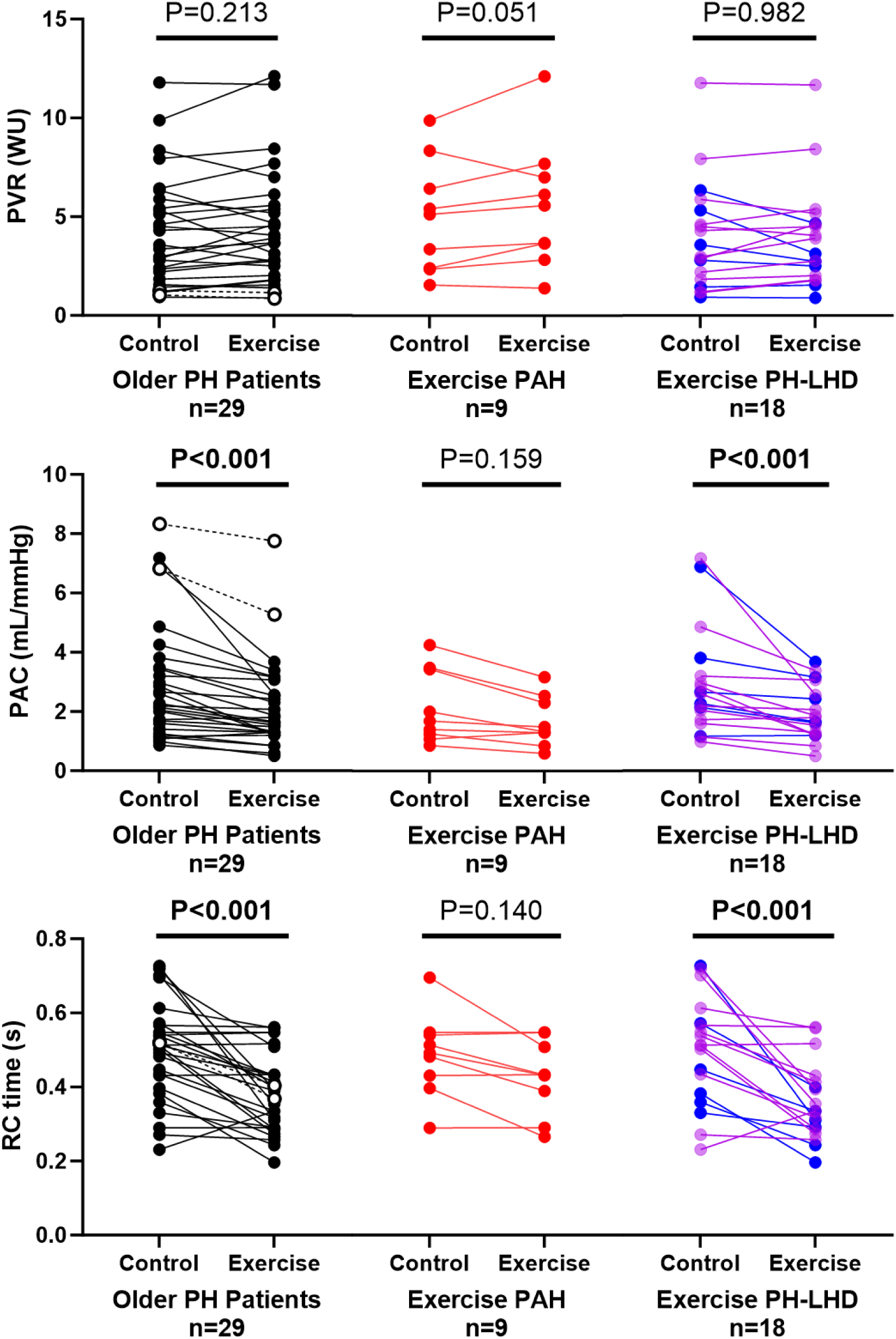
PVR, PAC, and RC time across the Exercise groupings at control and exercise. Black represents the exercise PH groups, open circles represent patients with No PH at rest and exercise, red represents Exercise PAH, blue represents Exercise PH-LHD_isolated_, and purple represents Exercise PH-LHD_combined_.

### Predictive Value of Resting Hemodynamics for Exercise Hemodynamic Classification

We examined the relationships between resting hemodynamic variables, PAWP or TPG, and subsequent exercise pressure-flow slopes indicative of either the post-capillary contribution (ΔPAWP/ΔCO) or the pre-capillary contribution (ΔTPG/ΔCO) (Figure 5 and Table 5). Resting hemodynamic variables, including PAWP or TPG, were not related to the subsequent exercise ΔPAWP/ΔCO slope. Accordingly, ROC analysis showed that resting hemodynamics did not discriminate ΔPAWP/ΔCO slope ≤ vs >2WU (Figure 6 and Table 6).

**Figure 5.**
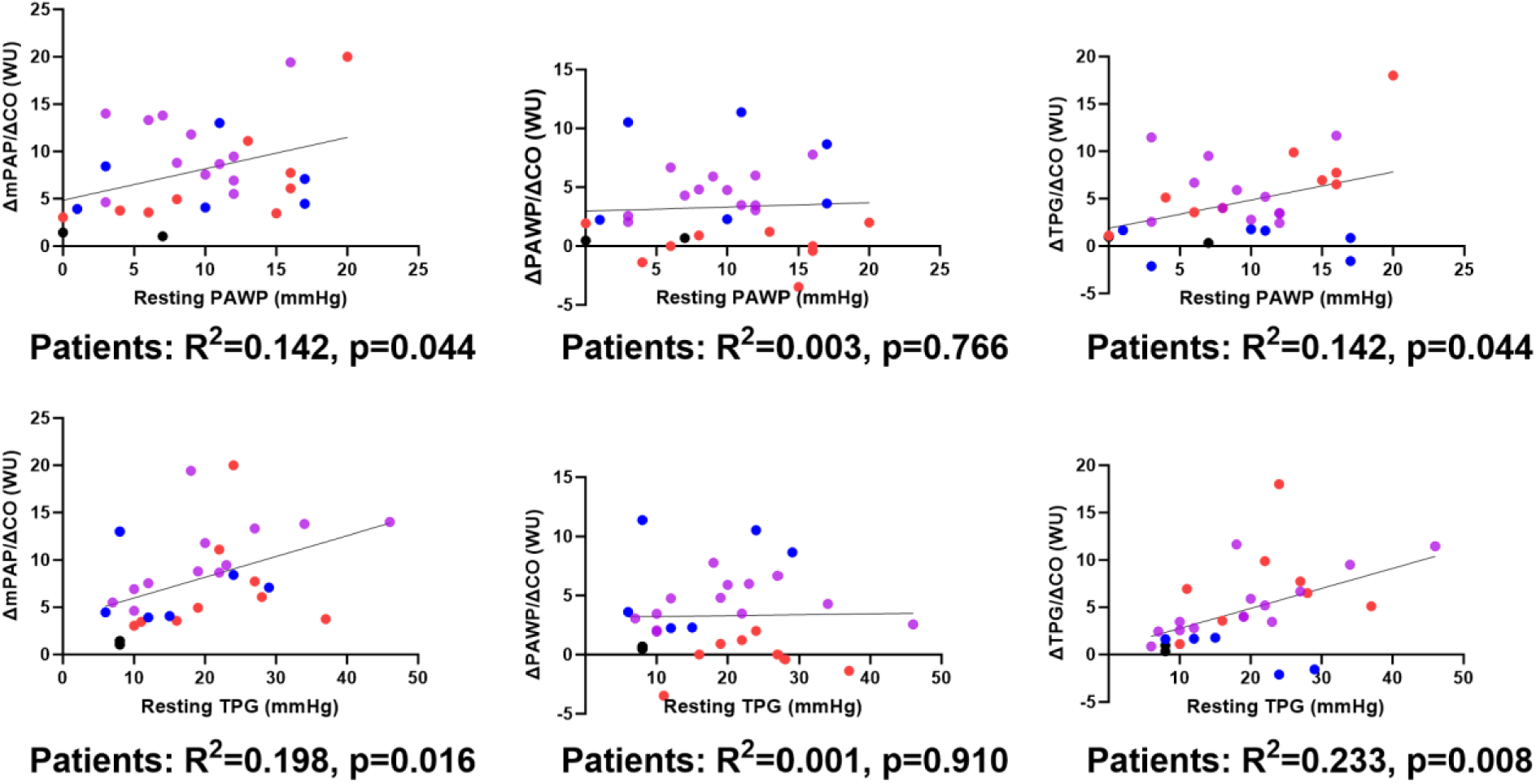
Linear regression of resting PAWP and TPG vs exercise pressure-flow slopes. Black circles represent No Ex PH, Red circles represent Exercise PAH, blue represents Exercise PH-LHD_isolated_, and purple represents Exercise PH-LHD_combined_.

**Figure 6.**
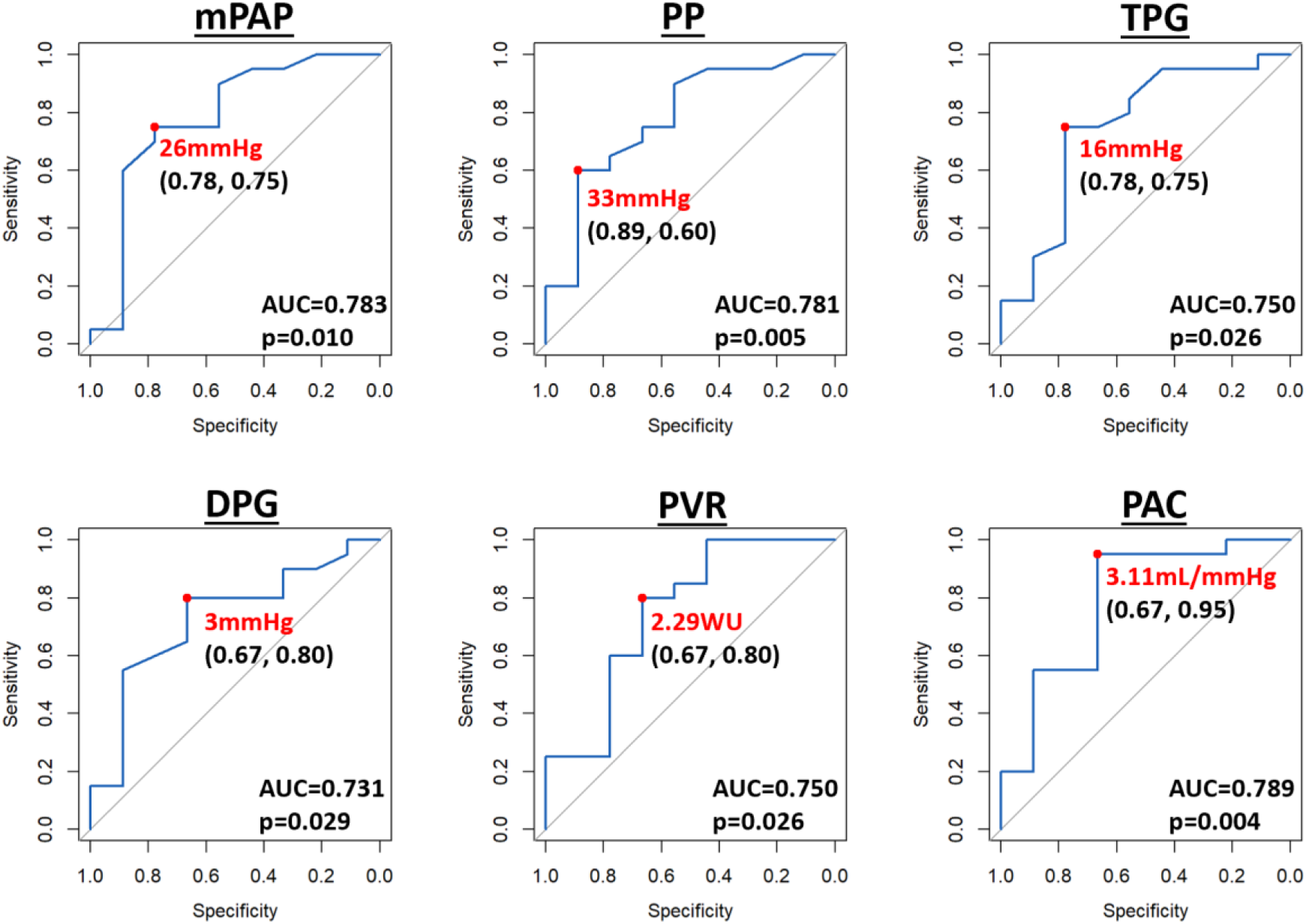
ROC of significant resting hemodynamics that have significant predictive ability to discriminate TPG/CO slope > vs ≤2WU in the older PH patient group. Red dot and text represent the optimal cut-off based on the Youden’s Index. In the label, x-coordinate represents specificity and y-coordinate represents the sensitivity.

**Table 5.** Linear Regression Analysis between resting hemodynamics and pressure-flow slopes in older PH patients.

| | ΔmPAP/ΔCO slope | | ΔPAWP/ΔCO slope | | ΔTPG/ΔCO slope | |
| --- | --- | --- | --- | --- | --- | --- |
|  | R<sup>2</sup> | P-value | R<sup>2</sup> | P-value | R<sup>2</sup> | P-value |
| HR (bpm) | 0.001 | 0.867 | 0.0006 | 0.897 | 0.003 | 0.771 |
| SV (mL) | 0.208 | **0.013** | 0.054 | 0.226 | 0.109 | 0.080 |
| RAP (mmHg) | 0.054 | 0.225 | 0.0004 | 0.922 | 0.061 | 0.198 |
| mPAP | 0.324 | **0.001** | 0.002 | 0.808 | 0.363 | **0.001** |
| PP | 0.092 | 0.109 | 0.010 | 0.601 | 0.177 | **0.023** |
| PAWP | 0.142 | **0.044** | 0.003 | 0.766 | 0.142 | **0.044** |
| TPG | 0.198 | **0.016** | 0.0005 | 0.910 | 0.233 | **0.008** |
| DPG | 0.148 | **0.040** | <0.002 | 0.997 | 0.187 | **0.019** |
| PVR (WU) | 0.292 | **0.0025** | 0.003 | 0.793 | 0.321 | **0.001** |
| PAC (mL/mmHg) | 0.339 | **0.0009** | 0.023 | 0.430 | 0.285 | **0.003** |
| RC time (s) | 0.114 | 0.074 | 0.055 | 0.219 | 0.037 | 0.314 |

AUC: area under the curve; 95% CI: confidence interval. Bolded p-values indicate P<0.05.

**Table 6.**
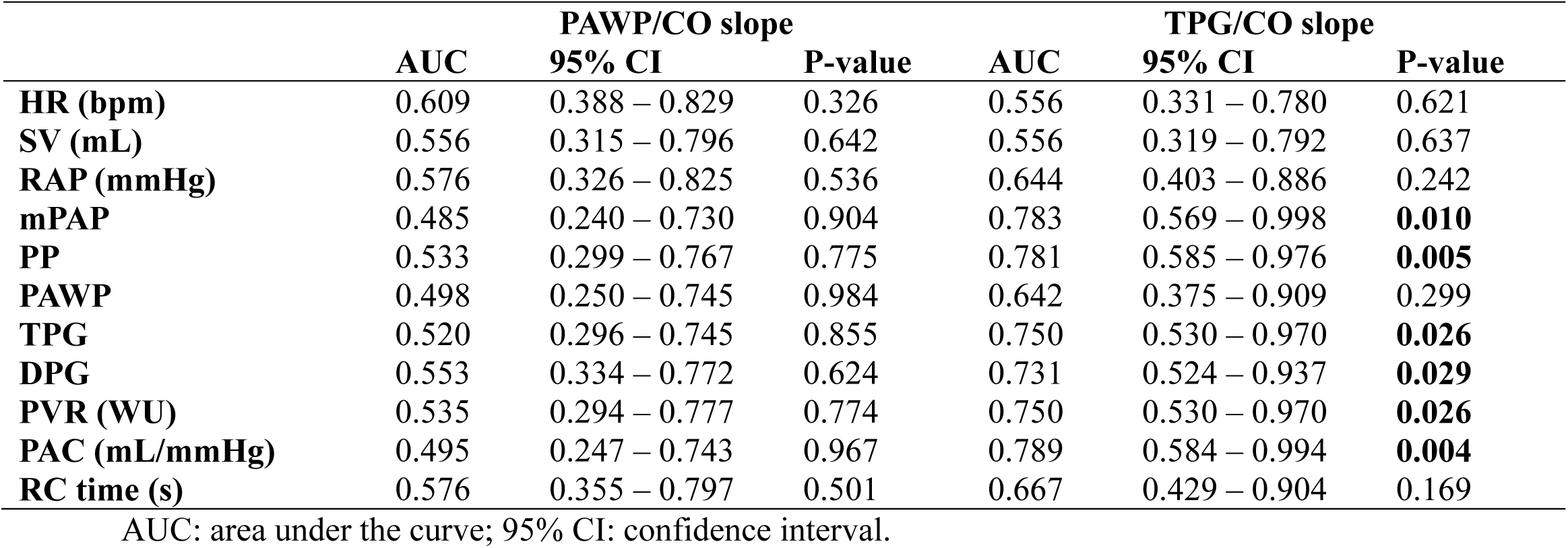
ROC analysis for the older PH patients demonstrating capability of resting hemodynamics to discriminate normal vs abnormal exercise pressure-flow slopes.

|  | PAWP/CO slope |  |  | TPG/CO slope |  |  |
| --- | --- | --- | --- | --- | --- | --- |
|  | AUC | 95% CI | P-value | AUC | 95% CI | P-value |
| **HR (bpm)** | 0.609 | 0.388 – 0.829 | 0.326 | 0.556 | 0.331 – 0.780 | 0.621 |
| **SV (mL)** | 0.556 | 0.315 – 0.796 | 0.642 | 0.556 | 0.319 – 0.792 | 0.637 |
| **RAP (mmHg)** | 0.576 | 0.326 – 0.825 | 0.536 | 0.644 | 0.403 – 0.886 | 0.242 |
| **mPAP** | 0.485 | 0.240 – 0.730 | 0.904 | 0.783 | 0.569 – 0.998 | **0.010** |
| **PP** | 0.533 | 0.299 – 0.767 | 0.775 | 0.781 | 0.585 – 0.976 | **0.005** |
| **PAWP** | 0.498 | 0.250 – 0.745 | 0.984 | 0.642 | 0.375 – 0.909 | 0.299 |
| **TPG** | 0.520 | 0.296 – 0.745 | 0.855 | 0.750 | 0.530 – 0.970 | **0.026** |
| **DPG** | 0.553 | 0.334 – 0.772 | 0.624 | 0.731 | 0.524 – 0.937 | **0.029** |
| **PVR (WU)** | 0.535 | 0.294 – 0.777 | 0.774 | 0.750 | 0.530 – 0.970 | **0.026** |
| **PAC (mL/mmHg)** | 0.495 | 0.247 – 0.743 | 0.967 | 0.789 | 0.584 – 0.994 | **0.004** |
| **RC time (s)** | 0.576 | 0.355 – 0.797 | 0.501 | 0.667 | 0.429 – 0.904 | 0.169 |

AUC: area under the curve; 95% CI: confidence interval.

In contrast, multiple resting hemodynamic variables, including both PAWP and TPG, were significantly related to exercise ΔTPG/ΔCO slope. In addition, resting mPAP, PP, DPG, PVR, and PAC were significantly related to the ΔTPG/ΔCO slope. Accordingly, ROC analysis demonstrated the area under the curve (AUC) reached statistical significance for resting mPAP, PP, TPG, DPG, PVR, and PAC to discriminate ΔTPG/ΔCO slope ≤ vs >2WU. Figure 6 illustrates the ROC analysis for these variables and their optimal cut-offs based on Youden’s index.

## DISCUSSION

An important conundrum for PH clinicians is reliance on resting hemodynamic criteria to differentiate PAH and PH-LHD amongst patients with left-heart risk factors. We tested the assumption that the likelihood that exercise would unmask post-capillary contributions to PH would be higher when resting PAWP was “borderline”. However, in the population of interest, we observed the relationship between resting PAWP and the final classification of Exercise PAH and PH-LHD were more complex. The predominant exercise classification was Exercise PH-LHD whether PAWP was “normal”, “borderline” or “high”. As such, resting PAWP did not predict Exercise PH-LHD classification which remained common even at lower resting PAWP ranges. At the same time, regardless of whether the exercise classification was Exercise PAH or Exercise PH-LHD, resultant peak exercise increases in mPAP were more strongly associated with pre-capillary contributions as measured by TPG. We also observed resting TPG did discriminate patients with abnormal pre-capillary pressure-flow slope behavior, regardless of resting PAWP values and whether final classification was Exercise PAH or PH-LHD. Overall, the highest exercise mPAP values observed were most strongly associated with pre-capillary contributions.

Our inclusion criteria were successful in prospectively recruiting patients typical of the challenges that PH clinicians face in regular practice, who harbour a high burden of left-heart risk factors. The prevalence of incident PH was high in this population, compared to exercise hemodynamic studies that predominantly included populations experiencing dyspnea of unknown origin (Bentley et al., 2020; Borlaug et al., 2010; Ho et al., 2020) and nearly all patients in the current study exhibited Exercise PH. Additionally, the majority satisfied the resting hemodynamic definition of PAH. However, we also observed a continuum of resting PAWP values (0 to 20mmHg) which did not suggest the presence of two distinct groups dichotomized by PAWP < or >15mmHg.

The pressure-flow slope criterion employed to define abnormal exercise post-capillary responses was based on findings from our own laboratory and others showing that the ΔPAWP/ΔCO does not exceed 2WU in healthy adults (Eisman et al., 2018; Esfandiari et al., 2017; Zeder et al., 2022). The majority of this population demonstrated a ΔPAWP/ΔCO slope >2WU with exercise; however, the relationships between the resting PAWP and the exercise responses were not straightforward. Exercise ΔPAWP/ΔCO slopes >2WU were observed in over 50% of patients with PAWP <12mmHg. Conversely, a final diagnosis of Exercise PAH was still observed in a proportion of patients grouped by PAWP ≥12mmHg or >15mmHg. Supine resting PAWP, even amongst proven heart failure with preserved ejection fraction, is dynamic based on volume status and treatment of decongestion (Zeder et al., 2024). Our study highlights the possibility that there may not be a lower cut-off for resting PAWP that reliably rules out dynamic left-heart abnormalities. Provocative testing to elicit latent PH-LHD may be better considered on the basis of clinical criteria rather than a single value threshold or borderline range for PAWP.

In this study, we also attempted to describe the balance of pre- and post-capillary contributions to mPAP and Exercise PH. Historical healthy control subjects were studied with a similar exercise protocol in our laboratory to illustrate the close relationship expected between mPAP and PAWP at rest and during exercise. In health, mPAP is primarily determined by left atrial pressure, which accounts for approximately 80% of the pressure in the PA (La Gerche et al., 2014). This relationship is reflective of a highly compliant and distensible pulmonary vasculature, that permits changes in left-heart pressures to be transmitted to the PA. In contrast, amongst our patient population, the relationship between mPAP and PAWP was highly variable and reflected disruption of normal pulmonary vascular function, irrespective of the spectrum of resting PAWP. As such, we employed the mPAP-PAWP difference, the TPG, as an index of pre-capillary contributions to PH. While TPG was very low across healthy subjects, amongst the patient population the relative strength of the association with mPAP was greater with TPG than PAWP, indicating the importance of pre-capillary contributions to PH regardless of whether the final classification was Exercise PAH or PH-LHD.

We employed pressure-flow slope analysis and ΔTPG/ΔCO >2WU, to further evaluate the relevance of pre-capillary pathophysiology, although there is less consensus regarding the appropriate thresholds for interpretation. The pressure-flow relationships suggested that pre-capillary pathophysiology contributed to exercise PH in approximately 70% of the study population. Notably, a proportion from each of the resting PAWP groups were ultimately classified as Exercise PAH, even from the PAWP ‘High’ zone, the middle and upper PAWP tertile. We further observed pre-capillary contributions to Exercise PH when exercise also disclosed ΔPAWP/ΔCO >2WU, which supports the notion that pre-and post-capillary pulmonary vascular mechanisms of disease are overlapping for many older PH patients. Moreover, increases in PAWP are known to augment pulsatile pulmonary vascular afterload relative to resistive afterload and associated shortening of RC time (Chemla et al., 2015; Tedford et al., 2012; Wright, Granton, et al., 2016). The current study upheld this construct; patients classified as Exercise PH-LHD, demonstrated significant lowering of PAC and shortening of RC time. This was in contrast to patients classified as Exercise PAH for whom RC time did not shorten or change significantly with exercise, also consistent with previous observations from our laboratory (Karvasarski et al., 2023; Tedford et al., 2012; Wright, Granton, et al., 2016).

From the clinical perspective, in a patient population with a high likelihood of incident PH and significant left-heart risk factors, an abnormal ΔPAWP/ΔCO slope could be elicited by exercise despite a resting PAWP below 12 mmHg and “borderline range”. Conversely, higher resting PAWP was observed in some of our older study population, but this did not rule out the development of pre-capillary pulmonary vascular disease. As such it is not surprising that within the range of PAWP studied, we could not identify a threshold resting PAWP value predicting the presence or absence of ΔPAWP/ΔCO >2WU. Conversely, any evidence of pre-capillary contributions to PH at rest (e.g., higher PVR, lower PAC, higher pulmonary PP, TPG or DPG) were associated with steeper ΔTPG/ΔCO slopes during exercise, regardless of resting PAWP. Exercise was also useful to fully expose pre-capillary abnormalities amongst patients eventually classified as Exercise PH-LHD, combined pre- and post capillary who generated exercise PA pressures similar to patients classified as Exercise PAH (Naeije et al., 2013).

There are limitations to our study that merit discussion. Our patient population may reflect the referral bias to our PH program; however, the challenge of older patients with PH is not unique to our center (Hoeper et al., 2013). We employed a 2-point method for calculation of pressure-flow slopes, which may be less robust than a multipoint assessment of both pressure and cardiac output (Ho et al., 2020). The ROC analysis suggests the ability for some resting hemodynamic measures to predict the pre-capillary contributions to PH; however, these findings require validation in an independent cohort.

## CONCLUSION

Our prospective experience shows that for older adults with risk factors for PH-LHD, in the presence of resting PAH hemodynamics, exercise may demonstrate a balance of pre- and post-capillary contributions to PH. From the current analysis, it is not clear that the indication for exercise to reveal post-capillary PH-LHD should be limited to patients with a PAWP between 12-15mmHg.

## Data Availability

The data supporting this study's findings are available from the corresponding author upon reasonable request.

## ACKNOWLEDGMENTS

The study was funded by Heart and Stroke Foundation of Canada Grant-in-Aid.

